# Prevalence of untreated HIV and HIV incidence among occupational groups in Rakai, Uganda: a population-based longitudinal study, 1999-2016

**DOI:** 10.1101/2022.06.21.22276714

**Authors:** Victor O. Popoola, Joseph Kagaayi, Joseph Ssekasanvu, Robert Ssekubugu, Grace Kigozi, Anthony Ndyanabo, Fred Nalugoda, Larry W. Chang, Tom Lutalo, Aaron A.R. Tobian, Donna Kabatesi, Stella Alamo, Lisa A. Mills, Godfrey Kigozi, Maria J. Wawer, John Santelli, Ronald H. Gray, Steven J Reynolds, David Serwadda, Justin Lessler, M. K. Grabowski

**Affiliations:** Department of Epidemiology, Johns Hopkins Bloomberg School of Public Health, Baltimore, Maryland; Rakai Health Sciences Program, Entebbe, Uganda; Makerere University School of Public Health, Kampala, Uganda; Division of Infectious Diseases, Department of Medicine, Johns Hopkins School of Medicine, Baltimore, Maryland, USA; Department of Pathology, Johns Hopkins School of Medicine, Baltimore, MD, USA; Division of Global HIV and TB, Centers for Disease Control and Prevention Uganda, Kampala Uganda; Department of Population and Family Health and Pediatrics, Columbia University, New York, NY, USA; Laboratory of Immunoregulation, Division of Intramural Research, National Institute for Allergy and Infectious Diseases, National Institutes of Health, Bethesda, Maryland, USA; Department of Epidemiology, UNC Gillings School of Global Public Health, Chapel Hill, NC, USA; Carolina Population Center, Chapel Hill, NC, USA

## Abstract

**Introduction:** Certain occupations have been associated with heightened risk of HIV acquisition and spread in sub-Saharan Africa, including bar work and transportation. However, data on changes in prevalence of untreated HIV infection and HIV incidence within occupations following rollout of antiretroviral therapy and voluntary medical male circumcision programs in 2004 are limited.

**Methods:** We evaluated 12 rounds of survey data collected between 1999-2016, from the Rakai Community Cohort Study, a population-based study of adolescents and adults 15-49 years in Uganda, to assess changes in the prevalence of untreated HIV infection and incidence by self-reported primary occupation. Adjusted prevalence risk ratios (adjPRR) for untreated HIV and incidence rate ratios for HIV incidence with 95% confidence intervals (CIs) were estimated using Poisson regression. Primary outcomes were stratified by gender and HIV incidence compared over three time periods (1999-2004; 2005-2011; 2011-2016) representing, respectively, the period prior to scale up of combined HIV prevention and treatment, the scale up period, and full implementation.

**Results:** 33,866 individuals, including 19,113 (56%) women participated. Of these participants, 17,840 women and 14,244 men who were HIV-negative at their first study visit contributed 57,912 and 49,403 person-years of follow-up, respectively. Agriculture was the most common occupation at all study visits, though its prevalence declined from 39 to 29% among men and from 61 to 40% among women between 1999 and 2016. Untreated HIV infection substantially declined between 1999 and 2016 across most occupational subgroups, including by 70% among men (12 to 4.2%; adjPRR=0.30; 95%CI:0.23-0.41) and by 78% among women working in agriculture (14.7 to 4.0%; adjPRR=0.22; 95%CI:0.18-0.27), along with increasing antiretroviral therapy coverage. Exceptions included men working in transportation and women working in tailoring/laundry services. HIV incidence declined in most occupations, but there were no reductions in incidence among female bar and restaurant workers or men working in transportation.

**Conclusion:** Untreated HIV infection and HIV incidence have declined in most occupational sub-groups in Rakai, Uganda. However, women working in bars and restaurants and men working in transportation continue to have relatively high burden of untreated HIV and HIV incidence, and as such should be considered key priority populations for targeted HIV programming.

## 1. INTRODUCTION

Mass scale-up of combination HIV treatment and prevention interventions (CHI) in sub-Saharan Africa has led to significant declines in HIV incidence [1–4]. However, rates of new HIV infection remain significantly above elimination thresholds in most countries [5, 6]. Demographic heterogeneities in population-level risk of HIV acquisition and onward transmission likely drive continued virus spread, but they remain poorly characterized. A detailed understanding of such heterogeneities may facilitate targeted control efforts leading to further declines in HIV incidence and, ultimately, disease elimination.

Decades-old data has established a person’s occupation as a salient risk factor for HIV acquisition in Africa. Occupations historically associated with increased HIV risk have included mining, bar work, truck driving, sex work, fishing, trading, and construction [3, 4, 7–10]. For example, a study of HIV risk in Uganda, conducted in 1992, prior to availability of antiretroviral therapy (ART), found that bar and restaurant work, trading, and truck and taxi driving were associated with three times higher odds of HIV acquisition compared to agricultural work [4]. In southern Africa, truck driving, factory work, and mining have been strongly linked to higher HIV burden [10–12]. While historical studies have provided useful insights into HIV risk by occupation, there are very limited data assessing key HIV outcomes within occupational subgroups since the widespread roll-out of CHI interventions in sub-Saharan Africa. Given that an individual’s occupation can be readily assessed in programmatic settings, understanding whether HIV burden currently varies across occupations may facilitate efficient targeting of interventions.

Here, we assess the extent to which occupation-specific prevalence of untreated HIV and HIV incidence have changed since the implementation of CHI programs, including ART and voluntary medical male circumcision (VMMC), using data from the Rakai Community Cohort Study (RCCS), a population-based study of HIV in south-central Uganda. We have previously measured trends in HIV prevalence and incidence in the RCCS and shown a 42% reduction in HIV incidence with ART rollout beginning in 2004 and VMMC scale-up beginning in 2007 [13]. However, it remains unclear whether or not untreated HIV prevalence and incidence declines have occurred uniformly across occupational subgroups in this population. We hypothesized that while the burdens of untreated HIV and HIV incidence have declined within all occupations, heterogeneities in HIV outcomes by occupation persist.

## 2. METHODS

### 2.1 Study population and procedures

The Rakai Community Cohort Study (RCCS) has been described in detail elsewhere [14]. In brief, the RCCS is conducted by the Rakai Health Sciences Program (RHSP) and is an open, population-based census and cohort study including consenting individuals aged 15-49 years across 40 communities. Individuals are followed at ∼18-month intervals. The RCCS conducts a household census to enumerate all individuals who are resident in the household, collecting data on gender, age, socioeconomic status, and global positioning system (GPS) location. The census is followed by a survey of all present consenting residents aged 15-49 years. Participant interviews provide data regarding sociodemographic characteristics, sexual behaviors, ART use, and whether they have had VMMC. Two attempts are made to contact individuals who are censused and eligible but who do not participate in the surveys.

To determine HIV status, venous blood samples are obtained for HIV testing. Prior to October 2011, HIV testing used enzyme immunoassays (EIAs) with confirmation via Western Blot. Subsequently, a field-validated parallel three-test rapid HIV testing algorithm was introduced with demonstrated high sensitivity (>99.5%) and specificity (>99.5%). All rapid test positives are confirmed by two EIAs, with Western Blot or PCR for discordant EIA results [15, 16].

In this study, we include data from 12 consecutive surveys conducted between April 6, 1999 and September 2, 2016 collected from 30 continuously surveyed RCCS communities. Participation rates among census eligible persons present in community at time of survey ranged from 74 to 98% (59-66% including those absent from community at time of survey) during this period [13]. The Rakai fishing communities were not included because data collection did not commence in that population until 2011. The 12 surveys are herein denoted as surveys 1 through 12: start and complete dates for each survey are included in Supplemental Table 1. This study was approved by the Research and Ethics Committee of the Uganda Virus Research Institute, the Ugandan Council of Science and Technology, and the Johns Hopkins School of Medicine Institutional Review Board.

### 2.2 Measurement and classification of participant occupation

Data were collected as self-reported primary and secondary occupations. Participants were asked “*What kind of work do you do, or what kind of activities keep you busy during an average day, whether you get money for them or not?*” In this study, we only analyzed primary occupation. There were 23 occupational subgroups that participants could select from on the questionnaire, including “*other.*” Individuals who listed “other” were asked to provide occupational details as a free-text response. Free-text responses were reviewed and re-assigned into pre-existing categories or new categories were created as needed. There were 41 self-reported primary occupations (Supplemental Tables 1a and 1b) which were aggregated into 15 occupational subgroups (Supplemental Table 2). Of these larger subgroups, 8 among men (agriculture, trading, student, construction, civil service, causal labor, mechanic, transportation) and 9 among women (agriculture, trading, student, bar/restaurant work, civil service, hairdressing, local crafts, tailoring/laundry, housekeeping) contained a median number of ≥ 50 observations per survey across all surveys (Supplemental Tables 3a and 3b).

**Table 1.** Characteristics of the study population at the baseline visit within each CHI calendar period by gender.

| Characteristics | Women |  |  | Men |  |  |
| --- | --- | --- | --- | --- | --- | --- |
|  | Visit 1 (1999-2000),<br>Pre-CHI baseline<br>N = 3,474 | Visit 6 (2005-2006),<br>Early-CHI baseline<br>N = 4,683 | Visit 10 (2011-2013),<br>Late-CHI baseline<br>N = 3,758 | Visit 1 (1999-2000),<br>pre-CHP baseline<br>N = 2,518 | Visit 6 (1999-2000),<br>early CHP baseline<br>N = 3,298 | Visit 10 (2011-2013),<br>late CHP baseline<br>N = 2,944 |
| **Age, Median (IQR)** | 25 (20-34) | 27 (22-34) | 28 (22, 35) | 26 (20, 33) | 28 (21, 35) | 27 (20, 36) |
| **Level of education** |  |  |  |  |  |  |
| None | 261 (7.5%) | 248 (5.3%) | 159 (4.2%) | 115 (4.6%) | 127 (3.9%) | 95 (3.2%) |
| Primary Education | 2312 (67%) | 2883 (62%) | 2082 (55%) | 1653 (66%) | 2050 (62%) | 1765 (60%) |
| Secondary Education | 779 (22%) | 1245 (27%) | 1206 (32%) | 545 (22%) | 837 (25%) | 814 (28%) |
| Tertiary Education | 119 (3.4%) | 297 (6.3%) | 279 (7.4%) | 203 (8.1%) | 259 (7.9%) | 214 (7.3%) |
| Missing | 3 (0.1%) | 10 (0.2%) | 32 (0.9%) | 2 (0.1%) | 25 (0.8%) | 56 (1.9%) |
| **Marital status** |  |  |  |  |  |  |
| Married | 2056 (59%) | 2811 (60%) | 2265 (60%) | 1418 (56%) | 1901 (58%) | 1524 (52%) |
| Never married | 781 (23%) | 1045 (22%) | 830 (22%) | 925 (37%) | 1153 (35%) | 1182 (40%) |
| Previously Married | 637 (18%) | 827 (18%) | 663 (18%) | 175 (6.9%) | 244 (7.4%) | 238 (8.1%) |
| **Religion** |  |  |  |  |  |  |
| None | 20 (0.6%) | 17 (0.4%) | 11 (0.3%) | 15 (0.6%) | 15 (0.5%) | 11 (0.4%) |
| Catholic | 2388 (69%) | 3061 (65%) | 2364 (63%) | 1713 (68%) | 2168 (66%) | 1867 (63%) |
| Protestant | 596 (17%) | 843 (18%) | 631 (17%) | 449 (18%) | 563 (17%) | 488 (17%) |
| Pentecostal | 43 (1.2%) | 123 (2.6%) | 162 (4.3%) | 32 (1.3%) | 93 (2.8%) | 131 (4.4%) |
| Muslim | 411 (12%) | 607 (13%) | 540 (14%) | 301 (12.0%) | 426 (13%) | 380 (13%) |
| Other | 13 (0.4%) | 20 (0.4%) | 18 (0.5%) | 6 (0.2%) | 8 (0.2%) | 11 (0.4%) |
| Missing | 3 (0.1%) | 12 (0.2%) | 32 (0.9%) | 2 (0.1%) | 25 (0.8%) | 56 (2%) |
| **Male circumcision** | - | - | - |  |  |  |
| Yes | - | - | - | 374 (15%) | 697 (21%) | 1359 (46%) |
| No | - | - | - | 2112 (84%) | 2516 (76%) | 1585 (54%) |
| **Untreated HIV prevalence** | 15.6% | 11.1% | 9.1% | 10.5% | 8.1% | 6.4% |

CHI=combination HIV intervention; Untreated HIV prevalence: prevalence of HIV-infected individuals who have not received antiretroviral therapy; IQR=interquartile range; Pre-CHI: 1999-2004; Early-CHI: 2005-2011; Late-CHI: 2012-2016

### 2.3 Primary and secondary outcomes

Our primary study outcomes were (1) untreated HIV infection and (2) incident HIV infection. We defined prevalent untreated HIV infection as HIV infection in an individual who did not self-report ART use. We have previously shown that self-reported ART use has high specificity (99%) and moderate sensitivity (77%) in this population [17]. Incident HIV infection was defined as a first HIV seropositive test result in a person with a prior seronegative test result. The unit of analysis for HIV incidence was person-years of follow-up between surveys, among persons who were initially HIV-negative and who contributed two consecutive survey visits or two visits with no more than one missing intervening survey. Incident infections were assumed to have occurred at the mid-point of the visit interval. Secondary outcomes included HIV prevalence overall, irrespective of self-reported ART use, and prevalence of self-reported ART use among HIV-positive persons.

### 2.4 Scale-up and measurement of Combination HIV Intervention coverage in Rakai

During the analysis period, antiretroviral therapy (ART) roll-out in Uganda, including Rakai, was phased as follows: in 2004, ART was offered to persons with a CD4-T-cell count of <250 cells/mm^3^; in 2011, the CD4 T-cell criterion was raised to <350; and in 2013, it was further increased to <500 and ART was also offered to all HIV-positive individuals, regardless of CD4 T-cell count, if they were pregnant, in a serodiscordant relationship, or self-identified as a sex worker or fisherfolk. Prevalence of self-reported ART use had risen to 69% of HIV positive individuals by 2016. In addition to ART, the RHSP has provided free VMMC since 2007 to adolescents and men age 13 years or older [13]. Prevalence of VMMC increased from 15% in 1999 to 59% by 2016 [13]. Impacts of universal HIV test and treat and pre-exposure prophylaxis impact were not assessed in this study as implementation of these programs occurred after the analysis period in 2017 and 2018, respectively.

To assess changes in HIV incidence by occupation over calendar time, we divided the study period into pre-CHI (1999-2004), early-CHI scale up (2005-2011), and mature-CHI (2012-2016) periods. Period-specific baselines were established as the first survey during each period, while the study baseline for individual participants was defined as their first survey during the study period.

### 2.5 Statistical analysis

Demographic characteristics of participants at period-specific baselines were summarized using descriptive statistics, including median and interquartile range for continuous variables and frequencies and percentages for categorical variables. The prevalence of each occupation was estimated as the number of participants in that occupation, expressed as a proportion of all participants surveyed and stratified by sex. Self-reported ART use among HIV positive participants was assessed during the early and mature-CHI periods and at the final study visit. Overall and untreated HIV prevalence were assessed at each of the 12 study visits and HIV incidence was estimated during the 11 inter-survey intervals over the 17-year analysis period. Primary and secondary outcomes were only assessed among the fifteen occupational subgroups (men, n=8; women, n=9). To evaluate changes in prevalence of untreated HIV infection and HIV incidence within occupational subgroups, we constructed log-binomial regression models to estimate prevalence risk ratios (PRR) and Poisson regression models to estimate incidence rate ratios (IRR). These IRRs and PRRs were adjusted for age and marital status. We calculated IRRs for HIV infection, comparing incidence rates during the pre-, early-, and late-CHI periods. All statistical analyses were performed in Stata version 15 and the R statistical software (Version 3.6).

## 3. RESULTS

### 3.1 Characteristics of study participants

Overall, 33,866 individuals, including 19,113 (56%) women participated, contributing a total of 102,759 person-visits. Of these participants, 17,840 women and 14,244 men who were HIV-negative at their first study visit contributed 57,912 and 49,403 person-years to the incidence cohort, respectively. Table 1 shows characteristics of the study population by sex at the first (baseline) study visit within the CHI periods. Among women, during the pre-CHI baseline visit, median age was 25.0 years (IQR: 20.0-34.0), 59% (2056/3474) were married, and the prevalence of untreated HIV was 16%. Median age at the late-CHI baseline visit for women was 28 years (IQR: 22.0-34.0), 60% (2265/3758) were married, and prevalence of untreated HIV infection was 9.1%. Among men, during the first pre-CHI baseline visit, median age was 26.0 years (IQR: 20.0-33.0), 56% (1418/2518) were married, 15% (374/2518) were circumcised, and the prevalence of untreated HIV infection was 8.1%. Median age at the late-CHI baseline visit for men was 27 years (IQR: 20.0-36.0), 52% (1524/2944) were married, 46% (1359/2944) were circumcised, and the prevalence of untreated HIV infection was 6.4%.

### 3.2 Population prevalence of occupations over calendar time

Figure 1 shows the proportion of participants in each occupational subgroup over calendar time stratified by sex. At the initial visit (1999-2000), the majority of women (61%) reported agriculture as their primary occupation. While agriculture remained the most commonly reported female occupation at the final visit (2015-16), its prevalence significantly declined to 40% (PRR 0.66, 95%CI: 0.62-0.69) (Figure 1, Supplemental Table 3a). Declines in agricultural work among women were accompanied by an increase in the average age within the occupation (Supplemental Figure 1) and were predominately offset by the proportion of women who reported working in trading (9.4% in 1999 vs.16% in 2016, PRR 1.7, 95%CI: 1.49-1.91) and being a student (7.3% vs. 14%, PRR 1.97, 95%CI: 1.72-2.27). Notably, no women or men reported sex work as a primary occupation, and very few people reported being unemployed (n<5 at all study visits; Supplemental Tables 1a-b).

**Figure 1.**
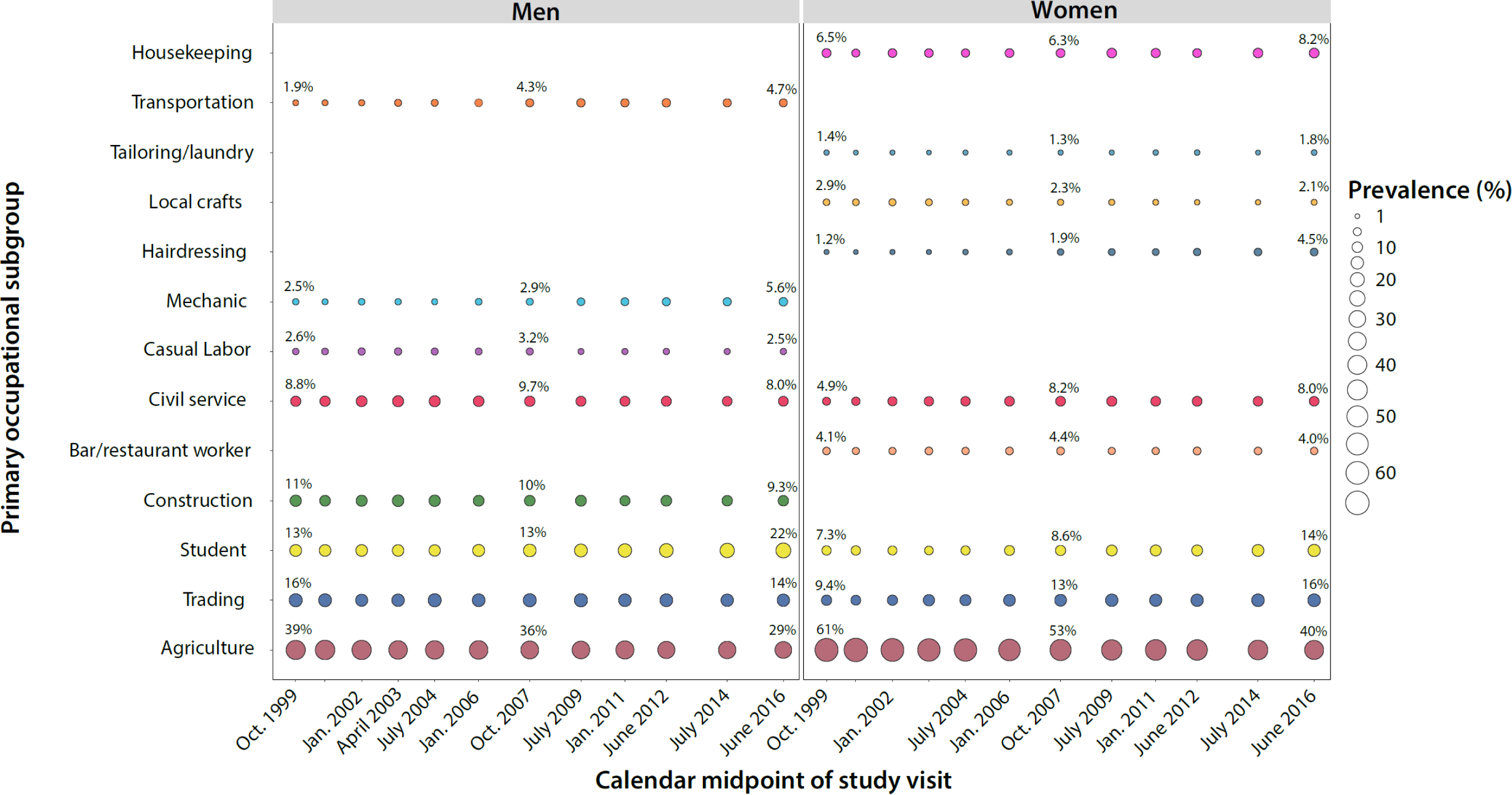
Prevalence of common primary occupational subgroups by sex in the Rakai Community Cohort Study, 1999-2016.

Men similarly reported agriculture and trading as their most common primary occupations (Figure 1, Supplemental Table 3b). Between the first (1999-2000) and last (2015-2016) study visit, there was a decrease in the proportion of male participants reporting agriculture (39% vs. 29%, PRR 0.74, 95%CI: 0.68-0.80), while a greater proportion reported being a student (13% vs. 22.0%, PRR 1.74, 95%CI: 1.53-1.96), mechanic (2.5% vs. 5.6%, PRR 2.29, 95%CI: 1.74-3.01), or working in transportation (1.9% vs. 4.7%, PRR 2.42, 95%CI: 1.78-3.28).

### 3.3 Trends in the prevalence of ART use and untreated HIV within occupations

The proportion of male and female HIV-positive participants self-reporting ART use increased over time among all occupational subgroups (Tables 2a-b). During the late-CHI period and at the final study visit, levels of ART use was highest among women working in agriculture and lowest among female students. ART use was statistically significantly lower among female traders (adjPRR=0.91; 0.83-0.98) and bar/restaurant workers (adjPRR=0.87; 95%CI: 0.78-0.97) compared to women working in agriculture during the late CHI-period. Among men, ART use was highest among those working in civil service over the entire analysis period. During the late CHI period, ART use was statistically significantly lower among men working in trading (adjPRR=0.91; 95%CI: 0.83-0.98) and male students (adjPRR: 0.59; 95%CI: 0.41-0.84).

**Table 2a:** Prevalence of self-reported ART use among HIV-positive women during the early and late-CHI periods and at the final study visit (Visit 12).

|  | Early – CHI (2004-2011)<br>N=3,352 |  |  | Late – CHI (2011-2016)<br>N=2,695 |  |  | Visit 12<br>N=1,059 |  |  |
| --- | --- | --- | --- | --- | --- | --- | --- | --- | --- |
|  | % self-reporting<br>ART (n/T) | PRR<br>(95% CI) | adjPRR<br>(95% CI) | % self-reporting<br>ART (n/T) | PRR<br>(95% CI) | adjPRR<br>(95% CI) | % self-reporting<br>ART (n/T) | PRR<br>(95% CI) | adjPRR<br>(95% CI) |
| Agriculture | 24.9<br>(432/1734) | Ref | Ref | 64.6<br>(811/1256) | Ref | Ref | 78.0<br>(375/481) | Ref | Ref |
| Trading | 25.5<br>(149/584) | 1.02<br>(0.87 - 1.20) | 1.04<br>(0.89 - 1.22) | 56.9<br>(302/531) | 0.88***<br>(0.81 - 0.96) | 0.91**<br>(0.83 - 0.98) | 68.2<br>(137/201) | 0.87**<br>(0.79 - 0.97) | 0.90**<br>(0.81 - 0.99) |
| Casual labor | - | - | - | - | - | - | - | - | - |
| Civil service | 19.5<br>(42/215) | 0.78*<br>(0.59 - 1.04) | 0.89<br>(0.68 - 1.17) | 58.2<br>(85/146) | 0.90<br>(0.78 - 1.04) | 0.93<br>(0.81 - 1.07) | 70.2<br>(40/57) | 0.90<br>(0.76 - 1.07) | 0.93<br>(0.78 - 1.11) |
| Student | 11.8<br>(2/17) | 0.47<br>(0.13 - 1.74) | 1.31<br>(0.35 - 4.92) | 38.0<br>(19/50) | 0.59***<br>(0.41 - 0.84) | 0.83<br>(0.58 - 1.20) | 53.3<br>(16/30) | 0.68**<br>(0.49 - 0.96) | 0.86<br>(0.60 - 1.22) |
| Bar/restaurant worker | 22.7<br>(65/287) | 0.91<br>(0.72 - 1.14) | 0.95<br>(0.76 - 1.20) | 55.9<br>(160/286) | 0.87**<br>(0.78 - 0.97) | 0.89**<br>(0.80 - 0.99) | 71.2<br>(79/111) | 0.91<br>(0.80 - 1.04) | 0.93<br>(0.82 - 1.06) |
| Local crafts | 12.8<br>(12/94) | 0.51**<br>(0.30 - 0.88) | 0.57**<br>(0.34 - 0.95) | 50.0<br>(32/64) | 0.77**<br>(0.60 - 0.99) | 0.82<br>(0.65 - 1.05) | 58.8<br>(20/34) | 0.76*<br>(0.57 - 1.00) | 0.79<br>(0.60 - 1.05) |
| Hairdressing | 21.0<br>(22/105) | 0.84<br>(0.58 - 1.23) | 1.13<br>(0.76 - 1.66) | 54.3<br>(51/94) | 0.84*<br>(0.70 - 1.02) | 0.95<br>(0.79 - 1.15) | 65.9<br>(27/41) | 0.85<br>(0.67 - 1.06) | 0.92<br>(0.74 - 1.15) |
| Tailoring/laundry | 23.7<br>(9/38) | 0.95<br>(0.53 - 1.69) | 1.07<br>(0.66 - 1.75) | 60.0<br>(18/30) | 0.93<br>(0.69 - 1.25) | 0.99<br>(0.75 - 1.31) | 75.0<br>(12/16) | 0.96<br>(0.72 - 1.28) | 1.01<br>(0.75 - 1.35) |
| Housekeeping | 12.4<br>(28/226) | 0.50***<br>(0.35 - 0.71) | 0.73*<br>(0.52 - 1.04) | 52.5<br>(94/179) | 0.81***<br>(0.70 - 0.94) | 0.96<br>(0.83 - 1.11) | 63.8<br>(44/69) | 0.82**<br>(0.68 - 0.98) | 0.90<br>(0.75 - 1.08) |
| Other occupations | 23.1<br>(12/52) | 0.93<br>(0.56 - 1.53) | 0.82<br>(0.50 - 1.33) | 66.1<br>(39/59) | 1.02<br>(0.85 - 1.24) | 1.04<br>(0.85 - 1.27) | 79.0<br>(15/19) | 1.01<br>(0.80 - 1.28) | 1.05<br>(0.82 - 1.36) |

**Table 2b:** Prevalence of self-reported ART use among HIV-positive women during the early and late-CHI periods and at the final study visit (Visit 12).

|  | Early – CHI (2004-2011)<br>N=1,702 |  |  | Late – CHI (2011-2016)<br>N=1,260 |  |  | Visit 12<br>N=464 |  |  |
| --- | --- | --- | --- | --- | --- | --- | --- | --- | --- |
|  | % self-reporting<br>ART (n/T) | PRR<br>(95% CI) | adjPRR<br>(95% CI) | % self-reporting<br>ART (n/T) | PRR<br>(95% CI) | adjPRR<br>(95% CI) | % self-reporting<br>ART (n/T) | PRR<br>(95% CI) | adjPRR<br>(95% CI) |
| Agriculture | 21.2<br>(140/661) | Ref | Ref | 54.4<br>(262/482) | Ref | Ref | 64.5<br>(118/183) | Ref | Ref |
| Construction | 7.8<br>(14/180) | 0.37***<br>(0.22 - 0.62) | 0.48***<br>(0.29 - 0.80) | 41.0<br>(64/156) | 0.76***<br>(0.62 - 0.93) | 0.86<br>(0.70 - 1.06) | 55.6<br>(30/54) | 0.86<br>(0.66 - 1.12) | 0.91<br>(0.69 - 1.19) |
| Trading | 14.9<br>(47/316) | 0.70**<br>(0.52 - 0.95) | 0.76*<br>(0.57 - 1.01) | 42.0<br>(94/224) | 0.77***<br>(0.65 - 0.92) | 0.80***<br>(0.67 - 0.94) | 57.8<br>(48/83) | 0.90<br>(0.73 - 1.11) | 0.91<br>(0.74 - 1.12) |
| Casual labor | 21.7<br>(15/69) | 1.03<br>(0.64 - 1.64) | 1.26<br>(0.79 - 2.02) | 36.9<br>(24/65) | 0.68**<br>(0.49 - 0.94) | 0.70**<br>(0.51 - 0.96) | 59.1<br>(13/22) | 0.92<br>(0.64 - 1.32) | 0.91<br>(0.64 - 1.29) |
| Civil service | 24.5<br>(34/139) | 1.16<br>(0.83 - 1.60) | 0.96<br>(0.69 - 1.34) | 60.6<br>(43/71) | 1.11<br>(0.91 - 1.37) | 1.03<br>(0.84 - 1.25) | 84.6<br>(22/26) | 1.31***<br>(1.08 - 1.60) | 1.21*<br>(1.00 - 1.48) |
| Student | 33.3 (4/12) | 1.57<br>(0.70 - 3.55) | 11.83***<br>(4.72 - 29.68) | 35.7 (5/14) | 0.66<br>(0.32 - 1.33) | 1.20<br>(0.56 - 2.55) | 50.0 (3/6) | 0.78<br>(0.35 - 1.74) | 1.21<br>(0.49 - 2.96) |
| Mechanic | 18.6<br>(11/59) | 0.88<br>(0.51 - 1.53) | 0.87<br>(0.54 - 1.40) | 43.3<br>(13/30) | 0.80<br>(0.53 - 1.21) | 0.80<br>(0.52 - 1.23) | 50.0 (7/14) | 0.78<br>(0.45 - 1.32) | 0.82<br>(0.50 - 1.34) |
| Transportation | 15.4<br>(18/117) | 0.73<br>(0.46 - 1.14) | 1.01<br>(0.67 - 1.52) | 42.9<br>(48/112) | 0.79**<br>(0.63 - 0.99) | 0.94<br>(0.76 - 1.17) | 52.6<br>(20/38) | 0.82<br>(0.59 - 1.13) | 0.94<br>(0.70 - 1.28) |
| Other occupations | 16.1<br>(24/149) | 0.76<br>(0.51 - 1.13) | 0.83<br>(0.57 - 1.21) | 50.9<br>(54/106) | 0.94<br>(0.76 - 1.15) | 1.01<br>(0.83 - 1.24) | 60.5<br>(23/38) | 0.94<br>(0.71 - 1.24) | 0.99<br>(0.76 - 1.29) |

Figures 2 and 3 show the prevalence of untreated HIV within occupational subgroups by gender at each of the 12 survey visits. Significant declines in the prevalence of untreated HIV were observed in nearly all occupational subgroups, irrespective of gender, with scale-up of ART use. Relative changes in untreated HIV between the initial and final visits are shown in Table 3 for each occupational subgroup by gender. The prevalence of untreated HIV significantly decreased within most occupations. For example, among women working in agriculture, prevalence of untreated HIV decreased from 15% to 4.0% (adjPRR 0.27; 95%CI:0.22-0.34) and among men, prevalence of untreated HIV decreased from 12% to 4% (adjPRR 0.27, 95%CI: 0.22-0.34).

**Figure 2a.**
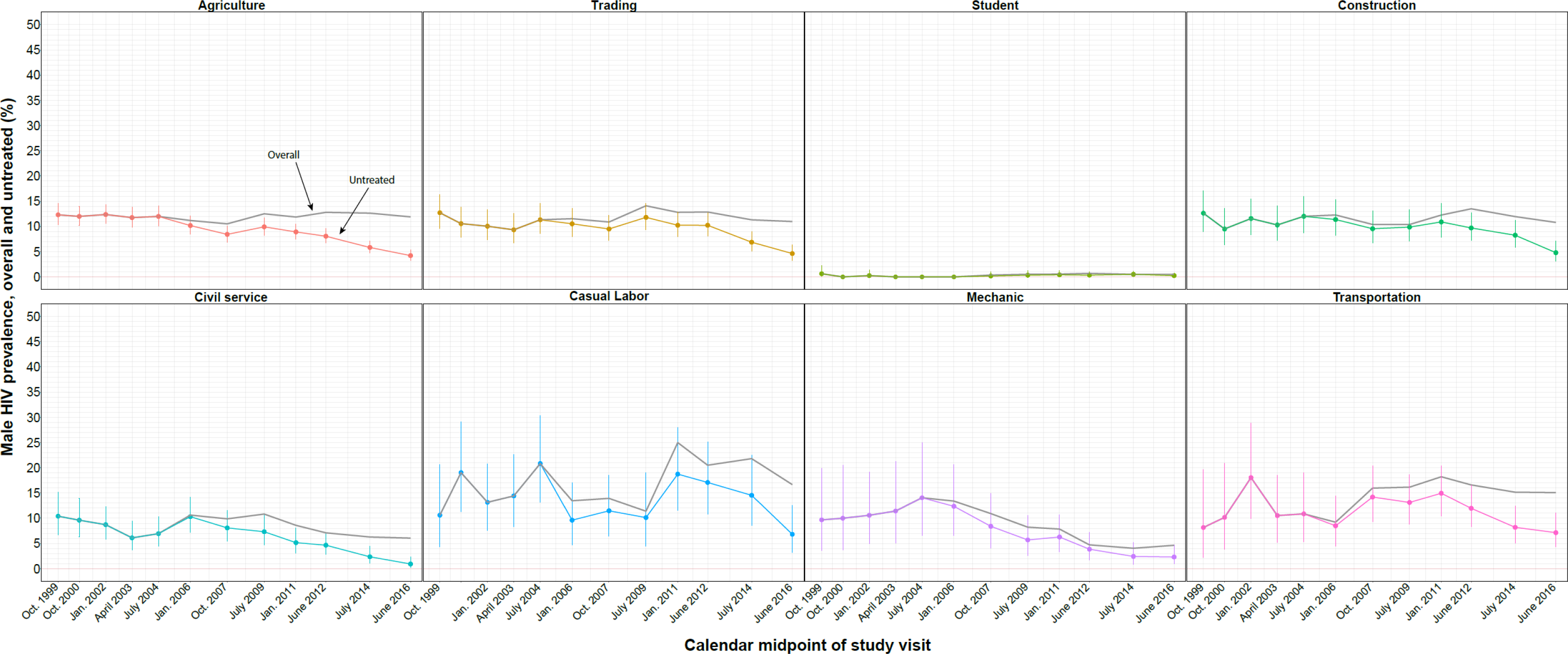
Trends in HIV prevalence overall (treated and untreated) among men by primary occupational subgroup in the Rakai Community Cohort Study, 1999-2016; Untreated prevalence with 95% confidence intervals are shown in color; overall HIV prevalence is shown in gray. Data are plotted at the calendar midpoint of the study visit.

**Figure 2b.**
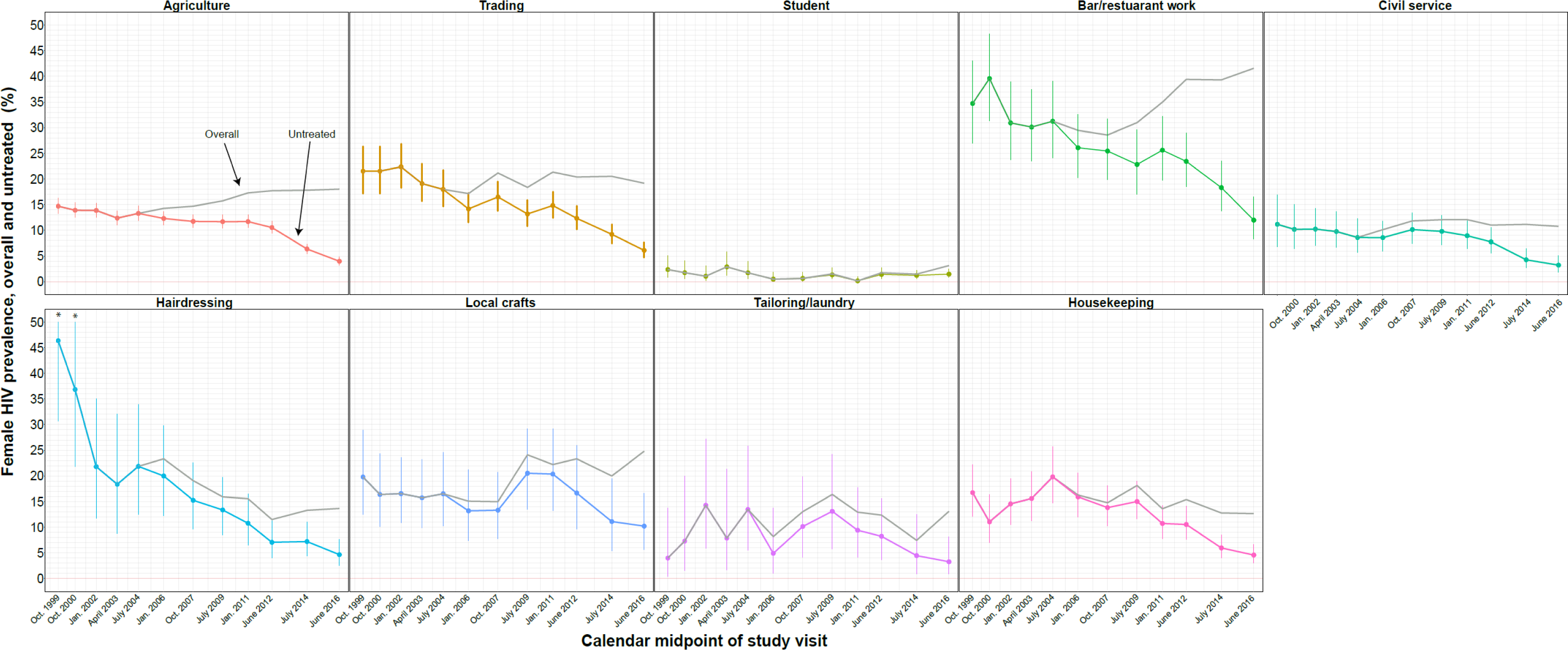
Trends in HIV prevalence overall (treated and untreated) among women by primary occupational subgroup in the Rakai Community Cohort Study, 1999-2016; Untreated prevalence with 95% confidence intervals are shown in color; overall HIV prevalence is shown in gray. Data are plotted at the calendar midpoint of the study visit. *Upper limit of 95%CI not shown.

**Table 3.** Change in prevalence of untreated HIV infection between visit 1 (1999-2000) and visit 12 (2015-2016)

| Occupational subgroup | Women<br>N = 10,121 |  |  |  |  | Men<br>N = 7,876 |  |  |  |  |
| --- | --- | --- | --- | --- | --- | --- | --- | --- | --- | --- |
|  | Visit 1<br>(1999-2000),<br>untreated<br>HIV<br>prevalence,<br>% (n/T)<br>n = 3474 | Visit 12<br>(2015-2016),<br>untreated<br>HIV<br>prevalence,<br>% (n/T)<br>n = 6647 | Unadjusted<br>PRR<br>(95% CI) | adjPRR*<br>(95% CI) | adjPRR<br>p-value | Visit 1 (1999-2000),<br>untreated<br>HIV<br>prevalence, %<br>(n/T)<br>n = 2,518 | Visit 12<br>(2015-2016),<br>untreated<br>HIV<br>prevalence,<br>% (n/T)<br>n = 5,358 | Unadjusted<br>PRR (95% CI) | adjPRR**<br>(95% CI) | adjPRR<br>p-value |
| Agriculture | 14.7<br>(313/2128) | 4.0<br>(106/2669) | **0.27 (0.22-0.23)** | **0.22 (0.18-0.27)** | **<0.001** | 12.3 (120/975) | 4.2 (65/1538) | **0.34 (0.26-0.46)** | **0.30 (0.23-0.41)** | **<0.001** |
| Construction | - | - | - | - | - | 12.6 (36/285) | 4.8 (24/500) | **0.38 (0.23-0.62)** | **0.27 (0.17-0.43)** | **<0.001** |
| Trading | 21.5<br>(70/325) | 6.1 (64/1048) | **0.28 (0.21-0.39)** | **0.09 (0.06-0.12)** | **<0.001** | 13.5 (51/401) | 4.6 (35/756) | **0.36 (0.24-0.55)** | **0.27 (0.17-0.43)** | **<0.001** |
| Casual labor | - | - | - | - | - | 10.6 (7/66) | 6.8 (9/132) | 0.64 (0.25-1.65) | 0.54 (0.22-1.35) | 0.185 |
| Civil service | 11.2<br>(19/170) | 3.2 (17/529) | **0.29 (0.15-0.54)** | **0.07 (0.04-0.15)** | **<0.001** | 10.4 (23/221) | 0.9 (4/429) | **0.09 (0.03-0.26)** | **0.09 (0.03-0.25)** | **<0.001** |
| Student | 2.4 (6/254) | 1.5 (14/959) | **0.62 (0.24-1.59)** | **0.11 (0.04-0.30)** | **<0.001** | 0.6 (2/319) | 0.3 (3/1178) | 0.41 (0.07-2.42) | 0.23 (0.04-1.19) | 0.079 |
| Mechanic | - | - | - | - | - | 9.7 (6/62) | 2.3 (7/302) | **0.24 (0.08-0.69)** | **0.22 (0.07-0.67)** | **0.007** |
| Transportation | - | - | - | - | - | 8.2 (4/49) | 7.1 (18/252) | 0.88 (0.31-2.47) | 0.90 (0.30-2.73) | 0.858 |
| Bar/Restaurant worker | 34.7<br>(50/144) | 12.0 (32/267) | **0.35 (0.23-0.51)** | **0.21 (0.14-0.31)** | **<0.001** | - | - | - | - | - |
| Local crafts | 19.8<br>(20/101) | 10.2 (14/137) | **0.52 (0.27-0.97)** | **0.29 (0.14-0.58)** | **<0.001** | - | - | - | - | - |
| Hairdressing | 46.3 (19/41) | 4.7 (14/300) | **0.10 (0.05-0.19)** | **0.01 (0.01-0.02)** | **<0.001** | - | - | - | - | - |
| Tailoring/laundry | 4.0 (2/50) | 3.3 (4/122) | 0.82 (0.16-4.33) | 0.25 (0.05-1.35) | 0.107 | - | - | - | - | - |
| Housekeeping | 16.7<br>(38/227) | 4.6 (25/545) | **0.27 (0.17-0.44)** | **0.11 (0.07-0.17)** | **<0.001** | - | - | - | - | - |
| Other occupations | 17.6 (6/34) | 5.6 (4/71) | 0.32 (0.10-1.06) | 0.28 (0.08-1.02) | 0.053 | 11.4 (16/140) | 5.5 (15/271) | **0.48 (0.25-0.95)** | **0.48 (0.25-0.96)** | **0.037** |
| All occupations | 15.6<br>(543/3474) | 4.4<br>(294/6647) | **0.28 (0.25-0.32)** | **0.27 (0.24-0.31)** | **<0.001** | 10.5<br>(265/2518) | 3.4<br>(180/5358) | **0.32 (0.27-0.38)** | **0.29 (0.24-0.35)** | **<0.001** |

PRR=prevalence risk ratios; adjPRR=adjusted prevalence risk; \*Models adjusted for age and marital status of study participants.

Women working in bars and restaurants had among the highest HIV burdens across all occupational subgroups. The prevalence of untreated HIV significantly declined among female bar and restaurant workers from a high of 35% in 1999-2000 to 12% by 2015-2016 (adjPRR 0.38; 95%CI: 0.25-0.58). However, these women still maintained a three-fold higher burden of untreated HIV, compared to women working in agriculture, at the final visit (12% vs. 4.0%). Women working in local crafts and in trading also continued to have a high prevalence of untreated HIV compared to women in agriculture at the final visit.

Men working in transportation did not have significantly higher HIV prevalence than other male occupations at the initial visit (Table 3). However, we observed no declines in untreated HIV in this population over the analysis period, and by the final visit, they had the highest prevalence of untreated HIV among all male occupations at 7.1%.

### 3.4 Changes in HIV incidence within occupations before and during scale-up of CHI programs

Table 4 shows HIV incidence by occupation, gender, and calendar time. In the early CHI period, HIV incidence rates ranged among the major occupations from 0.4 to 2.3 per 100 person-years among women, and from 0.1 to 1.8 per 100 person-years among men. Between the early and late CHI periods, HIV incidence declined among most occupational subgroups. For example, among those working in agriculture, HIV incidence declined by 67% among men (adjIRR 0.33, 95%CI: 0.21-0.54) and 38% among women (adjIRR 0.62; 95%CI: 0.45-0.86). HIV incidence trends in most other occupations showed a decline but were not statistically significant. While HIV incidence did not decline among students, incidence overall in this population was low. HIV incidence rates also did not decline among men working in transportation, and women working in bars and restaurants and local crafts.

**Table 4.** Incidence of HIV infection by occupational subgroup, sex, and CHI (combination HIV intervention) calendar period.

| Occupation | Women (N=17,840) |  |  |  |  |  |  |
| --- | --- | --- | --- | --- | --- | --- | --- |
|  | Incidence rate per 100 py (n/py) |  |  | IRR (95% CI) |  | adjIRR (95%CI) |  |
|  | Pre – CHI<br>(1999-2004) | Early – CHI<br>(2004-2011) | Late – CHI<br>(2011-2016) | Early – CHI vs.<br>Pre-CHI (ref) | Late – CHI vs. Pre-CHI (ref) | Early – CHI vs.<br>Pre-CHI (ref) | Late – CHI vs.<br>Pre-CHI (ref) |
| Agriculture | 1.1 (97/8490) | 0.9 (129/13785) | 0.7 (62/9515) | 0.82 (0.63 – 1.07) | **0.57\* (0.41 - 0.79)** | 0.87 (0.67 – 1.13) | **0.62\* (0.45 - 0.86)** |
| Bar/restaurant worker | 1.1 (4/365) | 2.1 (17/813) | 2.0 (12/605) | 1.91 (0.64 – 5.69) | 1.81 (0.58 – 5.66) | 2.13 (0.72 – 6.36) | 2.79 (0.85 - 9.19) |
| Trading | 1.4 (17/1222) | 1.4 (49/3474) | 0.7 (23/3117) | 1.01 (0.58 - 1.77) | **0.53\* (0.28 – 1.00)** | 1.19 (0.69 – 2.06) | 0.72 (0.38 - 1.37) |
| Hairdressing | 2.3 (3/133) | 1.6 (10/624) | 0.8 (6/774) | 0.71 (0.19 – 2.62) | 0.34 (0.08 – 1.39) | 0.73 (0.20 - 2.68) | 0.36 (0.09 – 1.43) |
| Civil service | 1.1 (9/851) | 0.6 (14/2399) | 0.3 (5/1849) | 0.55 (0.24 - 1.28) | **0.26\* (0.09 – 0.77)** | 0.76 (0.31 - 1.86) | 0.49 (0.13 - 1.78) |
| Student | 0.3 (2/623) | 0.4 (6/1433) | 0.7 (14/2059) | 1.30 (0.26 – 6.48) | 2.12 (0.48 - 9.35) | 1.23 (0.25 – 6.11) | 1.93 (0.44 – 8.36) |
| Housekeeping | 1.2 (6/496) | 1.1 (15/1388) | 0.9 (12/1278) | 0.89 (0.35-2.32) | 0.78 (0.29-2.08) | 1.0 (0.37-2.67) | 0.89 (0.30-2.59) |
| Local crafts | 1.3 (5/387) | 2.3 (12/518) | 1.6 (5/308) | 1.79 (0.63 – 5.13) | 1.25 (0.36 – 4.38) | 1.88 (0.65 – 5.44) | 1.36 (0.38 – 4.91) |
| Tailoring/laundry | 1.7 (2/121) | 1.4 (5/355) | 1.1 (3/261) | 0.86 (0.16-4.47) | 0.70 (0.12-4.23) | 0.94 (0.17-5.10) | 0.85 (0.15-4.84) |
| Other occupations | 1.1 (1/93) | 0.0 (0/169) | 1.2 (5/406) | - | 1.14 (0.13 - 9.96) | - | 1.41 (0.16-12.48) |
| All occupations | 1.1 (146/12781) | 1.0 (257/24958) | 0.7 (147/20173) | 0.90 (0.74 - 1.11) | **0.64\* (0.51 - 0.80)** | 0.93 (0.76-1.14) | **0.66\* (0.53-0.83)** |
|  | Men (N=14,244 ) |  |  |  |  |  |  |
|  | Incidence rate per 100 py (n/py) |  |  | IRR (95% CI) |  | adjIRR (95%CI) |  |
|  | Pre – CHI<br>(1999-2004) | Early – CHI<br>(2004-2011) | Late – CHI<br>(2011-2016) | Early – CHI vs.<br>Pre-CHI (ref) | Late – CHI vs Pre-CHI (ref) | Early – CHI vs.<br>Pre-CHI (ref) | Late – CHI vs<br>Pre-CHI (ref) |
| Agriculture | 1.4 (53/3894) | 0.8 (62/8046) | 0.4 (25/5834) | **0.57\* (0.39 - 0.82)** | **0.32\* (0.20 - 0.51)** | **0.58\* (0.40 - 0.84)** | **0.33\* (0.21 - 0.54)** |
| Construction | 1.6 (18/1128) | 1.2 (28/2322) | 0.5 (9/1690) | 0.76 (0.42 - 1.37) | **0.33\* (0.15 - 0.75)** | 0.79 (0.42 - 1.47) | **0.35\* (0.15 - 0.83)** |
| Trading | 0.8 (13/1541) | 0.7 (26/3613) | 0.5 (15/2746) | 0.85 (0.44 - 1.67) | 0.65 (0.31 - 1.37) | 0.89 (0.46 - 1.70) | 0.69 (0.33 - 1.45) |
| Casual labor | 1.2 (3/259) | 1.6 (7/431) | 0.6 (2/322) | 1.40 (0.36 - 5.49) | 0.54 (0.09 - 3.25) | 1.67 (0.42 - 6.66) | 0.68 (0.11 - 4.30) |
| Civil service | 0.7 (7/981) | 0.6 (12/2121) | 0.4 (6/1673) | 0.79 (0.31 - 2.02) | 0.50 (0.17 - 1.50) | 0.78 (0.31 - 2.00) | 0.49 (0.16 - 1.51) |
| Student | 0.1 (1/955) | 0.05 (1/1905) | 0.1 (3/2840) | 0.50 (0.03 - 8.03) | 1.01 (0.11 - 9.71) | 0.51 (0.03 - 8.23) | 0.44 (0.04 - 4.86) |
| Mechanic | 0.0 (0/228) | 1.0 (8/780) | 0.4 (4/901) | - | - | - | - |
| Transportation | 1.4 (4/287) | 1.8 (21/1181) | 1.2 (12/964) | 1.28 (0.43 - 3.75) | 0.89 (0.29 - 2.80) | 1.33 (0.45 - 3.91) | 1.10 (0.35 - 3.50) |
| Other occupations | 1.8 (10/546) | 1.9 (21/1099) | 0.9 (10/1115) | 1.04 (0.49 - 2.23) | 0.49 (0.20 - 1.19) | 1.06 (0.50 - 2.26) | 0.50 (0.21 - 1.23) |
| All occupations | 1.1 (109/9821) | 0.9 (186/21498) | 0.5 (86/18085) | **0.78\* (0.62 - 0.99)** | **0.43\* (0.32 - 0.57)** | **0.79\* (0.62 - 1.0)** | **0.44\* (0.33 - 0.59)** |

py=person years; IRR=incidence rate ratio; adjIRR=adjusted incidence rate ratio for age and marital status; IRR not presented for other occupations (women, Early-CHI) and mechanic (men) because there were no cases in the numerator and denominator respectively; CHI=combination HIV intervention \*p<0.05

Table 5 shows the adjusted relative risk of HIV acquisition by occupation during the late CHI period. Compared to women working in agriculture, female bar and restaurant workers had a three-fold higher HIV incidence (adjIRR 2.88, 95% CI 1.51-5.49, p = 0.001). Men working in transportation also had increased HIV incidence compared to agricultural workers (adjIRR 2.75, 95% CI 1.37-5.50). Regardless of sex, students had a significantly lower risk of HIV acquisition compared to persons working in agriculture (men: adjIRR 0.19, 95% CI 0.05-0.73; women: adjIRR 0.36, 95% CI 0.18-0.72).

**Table 5.** Adjusted incidence rate ratios of HIV infection comparing all occupations vs. agriculture during the late-CHI period

| Occupation | Women |  | Men |  |
| --- | --- | --- | --- | --- |
|  | adjIRR (95% CI) | p-value | adjIRR (95% CI) | p-value |
| Agriculture | Ref. | Reference | Reference | Reference |
| Construction | - | - | 1.16 (0.54-2.48) | 0.707 |
| Trading | 1.1 (0.66-1.73) | 0.800 | 1.25 (0.66-2.37) | 0.501 |
| Casual labor | - | - | 1.45 (0.33-6.39) | 0.623 |
| Civil service | 0.40 (0.16-1.00) | 0.051 | 0.87 (0.36-2.13) | 0.767 |
| Student | **0.36 (0.18-0.72)** | **0.004** | **0.19 (0.05-0.73)** | **0.015** |
| Mechanic | - | - | 0.91 (0.30-2.83) | 0.875 |
| Transportation | - | - | **2.75 (1.37-5.50)** | **0.004** |
| Bar/restaurant worker | **2.88 (1.51-5.49)** | **0.001** | - | - |
| Local crafts | 1.96 (0.76-5.06) | 0.164 | - | - |
| Hairdressing | 0.76 (0.32-1.79) | 0.523 | - | - |
| Tailoring/laundry | 1.33 (0.40-4.42) | 0.640 | - | - |
| Housekeeping | 1.08 (0.57-2.04) | 0.819 | - | - |

adjIRR=adjusted incidence rate ratio for age and marital status
Occupations where the median number of observations per visit was <50 were excluded from the analysis.

## 4. DISCUSSION

In this population-based study, we observed declining prevalence of untreated HIV and HIV incidence among most occupational subgroups with the scale up of CHI programs in Uganda. Among men and women working in agriculture, the most common primary occupation, prevalence of untreated HIV and HIV incidence declined by more than two-thirds. However, this was not always the case for other occupations. While women working in bars and restaurants made up a small proportion of the overall population, they had among the highest burdens of untreated HIV prior to CHI scale-up, with no declines in HIV incidence over the analysis period. We also found no significant reduction in HIV incidence among male transportation workers. Moreover, both female bar and restaurant workers and male transportation workers had the highest prevalence of untreated HIV at the final study visit. HIV incidence rates among women reporting student and crafting as primary occupations also showed no decrease following CHI scale-up, although students had a very low HIV burden overall. Taken together, these results suggest that members of traditionally high-risk occupations continue to experience elevated rates of HIV incidence and remain challenging to engage in HIV programs.

Other studies have reported high HIV prevalence among female bar workers in sub-Saharan Africa [18, 19]. In this study, HIV prevalence among female bar and restaurant workers exceeded 40% with rising prevalence in recent years. While the prevalence of untreated HIV significantly declined in this population, it was three times higher than among women working in agriculture at the final study visit. Female bar work also has been associated with a heightened risk of HIV incidence in prior studies [20, 21]. The high burden of HIV among these women has been linked to elevated rates of transactional sex, alcohol use, and mobility [21–23]. In a systematic review of socio-demographic characteristics and risk factors for HIV among female bar workers, high rural-to-urban mobility, transactional sex, and inconsistent condom use were common and associated with financial needs and social marginalization [21]. Our results underscore that female bar and restaurant workers continue to be a priority population for African HIV treatment and prevention programs. While key population-based programs in Africa include female sex workers, and many female bar workers are engaged in sex work, not all women working in bars and restaurants at high risk of HIV classify themselves as sex workers [21]. Multi-level, social influence, and structural HIV prevention interventions targeting alcohol-serving establishments, including enhanced sexually transmitted infection clinic services, portable health services, and peer education, have been shown to be effective in settings outside Africa, for reducing HIV risk and increasing treatment uptake [24, 25].

Prior research has shown that men working in transportation are highly mobile and often engage in transactional sex [26–28]. We found that the prevalence of untreated HIV did not significantly decline in this occupational sub-group with the availability of CHI. Prior research has linked male transportation workers, including truck drivers, to higher risk of HIV transmission [26], and has shown that men working in this occupation frequently engage with sex workers and women working in bars and restaurants [27, 29]. Supplies of free condoms, roadside clinics, and free HIV testing services at truck stops are some HIV prevention interventions that have targeted male transportation workers [10, 29]; however, levels of awareness and uptake of such services in this population has been low [10, 30].

Adolescent girls and young women aged 15 to 24 years have a disproportionately high risk of HIV acquisition in Africa [31–33], but HIV risk was significantly lower among young people who list their occupation as “student” and who have higher education attainment, regardless of sex [34–37]. During the study period, HIV prevalence declined in female students by nearly 90%. Incidence of HIV remained stable for both male and female students, but compared to those in agriculture, students of both sexes had lower HIV incidence during the late-CHI period. Research from South Africa has shown that students tend to have smaller sexual networks and are less likely to report high-risk sexual behaviors compared to those not in school [35]. Lower HIV incidence and prevalence among female students has also been attributed to avoiding the consequences of unprotected sex and increased self-efficacy for negotiating safer sex with their partners [38]. Interventions that increase school enrollment of adolescent girls and young women, such as PEPFAR’s Determined, Resilient, Empowered, AIDS-free, Mentored, and Safe (DREAMS) program, may decrease sexual initiation, high-risk sexual behavior, and HIV risk [31]. Since the onset of the COVID-19 pandemic in Uganda during the spring of 2020, schools have remained fully or partially closed. Given the strong protective effects of schooling on HIV acquisition, understanding the extent to which school closures impact HIV and other reproductive health outcomes, such as unplanned pregnancy, is an urgent public health priority.

Earlier studies have established migration and mobility as a key risk factor for HIV acquisition and transmission [22, 39, 40]. Overall, we found that the occupations which tend to have high mobility also had higher prevalence of untreated HIV and HIV incidence despite CHI scale-up. Both female bar and restaurant work and male transportation work are associated with increased mobility as well as high-risk sexual behaviors, including concurrent sexual partnerships and inconsistent condom use [27, 28, 41]. Specialized service-delivery tailored to mobile populations include the implementation of client-managed groups, adherence clubs, community drug distribution points, and multi-month prescriptions to reduce HIV burden in these populations [42–44].

The shifting distribution of the occupational makeup in our study population away from agriculture likely reflects the increasing urbanization happening across the African continent [45]. Little data exists on the impact of urbanization on HIV transmission; however, in sub-Saharan Africa, HIV prevalence and incidence have been reported to be higher in urban than in rural centers [46, 47]. This has been attributed to factors such as relative affluence in urban centers, increased social interaction, and higher-risk behaviors such as transactional sex and concurrent sexual partnerships [48–50]. More research is needed to elucidate the impact of increasing urbanization on HIV transmission within African populations.

Our study has important limitations. Occupation and ART use were self-reported and may be subject to bias. However, we have previously shown that self-reported ART has high specificity and moderate sensitivity in this same study population [17]. Sex work in Uganda is criminalized and was likely underreported in our survey [4]. Key population-focused HIV prevention programs began in this region in 2017, after the time of the analysis, and so their impact cannot be assessed. However, our findings do support their focus on female sex workers. Our data suggest that other population sub-groups (beyond the currently defined key and priority population groups in Uganda) may merit additional HIV prevention focus, including bar and restaurant workers and short/medium distance transportation workers. While the longitudinal nature of this study is a strength, analysis of incident HIV infections was limited by a small number of events in some occupational subgroups, which may have obscured trends. Lastly, our results may not be generalizable to other populations, particularly in urban African settings where the nature of the association between occupation and HIV risk may differ.

## 5. CONCLUSIONS

In summary, prevalence of untreated HIV infection and HIV incidence declined in most occupational subgroups following the mass scale-up of HIV prevention and treatment interventions in rural southern Uganda. However, HIV burden remained relatively high in the traditionally high-risk occupations of male transportation and female bar and restaurant work. HIV programs that meet the unique needs of these high-risk populations, which tend to be more mobile with higher levels of HIV-associated risk behaviors, may help achieve HIV epidemic control.

## Data Availability

All data produced in the present study are available upon reasonable request to the authors

## ACKNOWLEGEMENTS

We thank the RCCS participants and many staff and investigators who have made this study possible over the years. Additionally, we thank the personnel at the Office of Cyberinfrastructure and Computational Biology at the National Institute of Allergy and Infectious Diseases for data management support.

## FUNDING

This study was supported by the National Institute of Allergy and Infectious Diseases (grants R01AI110324, U01AI100031, U01AI075115, R01AI102939, R01AI14333, R01AI155080, and K01AI125086-01), the National Institute of Mental Health (grants R01MH107275 and R01MH105313), the Eunice Kennedy Shriver National Institute of Child Health and Human Development (grants R01HD070769 and R01HD050180), the Division of Intramural Research of the National Institute for Allergy and Infectious Diseases, the Johns Hopkins University Center for AIDS Research (grant P30AI094189), and the President’s Emergency Plan for AIDS Relief through the Centers for Disease Control and Prevention (grant NU2GGH000817). The findings and conclusions in this article are those of the authors and do not necessarily represent the official position of the funding agencies.

## CONFLICTS OF INTEREST

Drs. Wawer and Gray are paid consultants to the Rakai Health Sciences Program and serves on its Board of Directors. These arrangements have been reviewed and approved by the Johns Hopkins University in accordance with its conflict of interest policies.

---

**Supplemental Figure 1.**
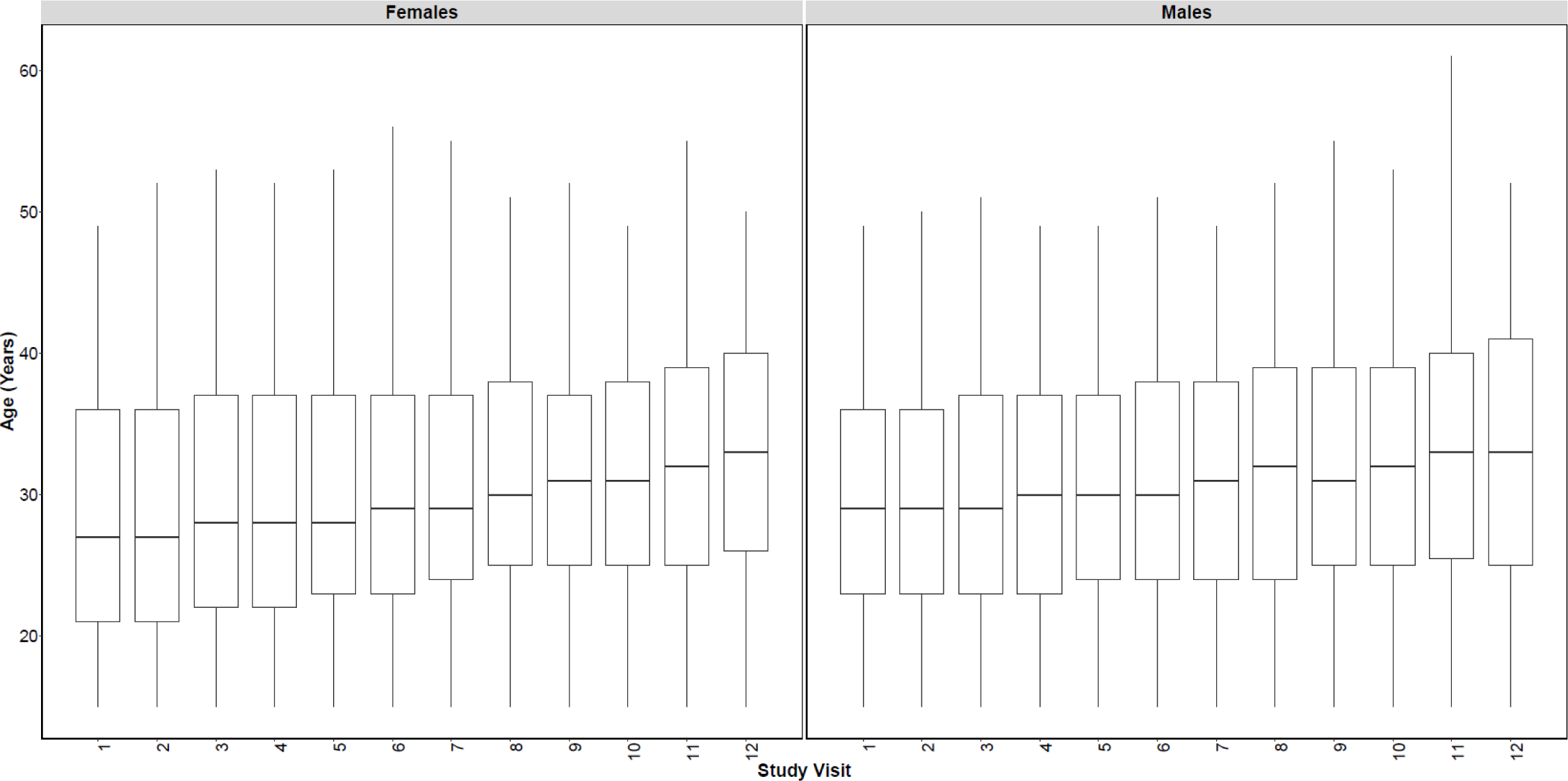
Boxplots of age in years at each study visit, among RCCS agricultural workers. The bottom and top of the boxes represent the 25th and 75th percentiles of the data (interquartile range), respectively. The solid black line in the middle of the box represents the median (50th percentile) and the lines represent values beyond the middle 50% of the data.

**Supplemental Table 1a.**
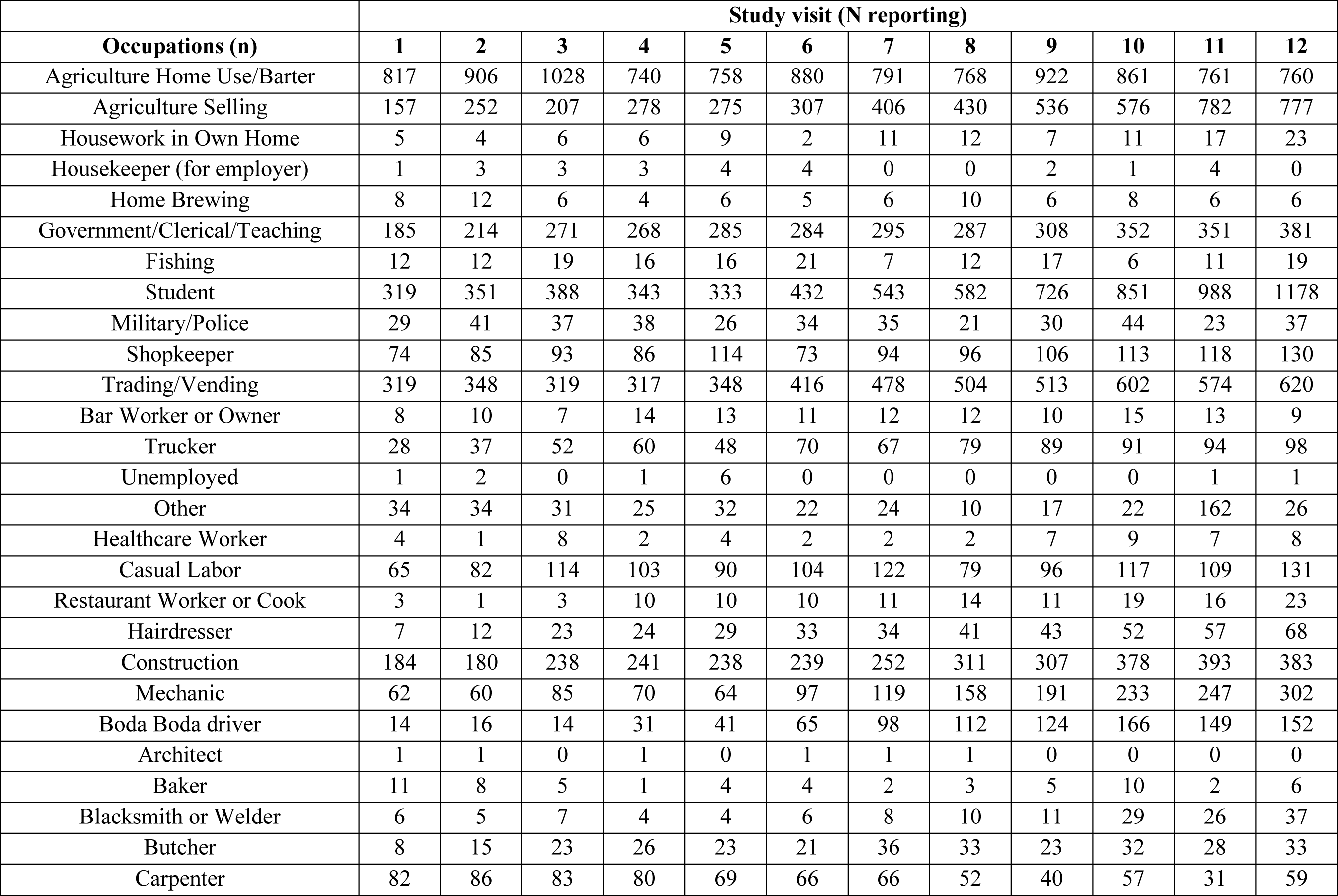
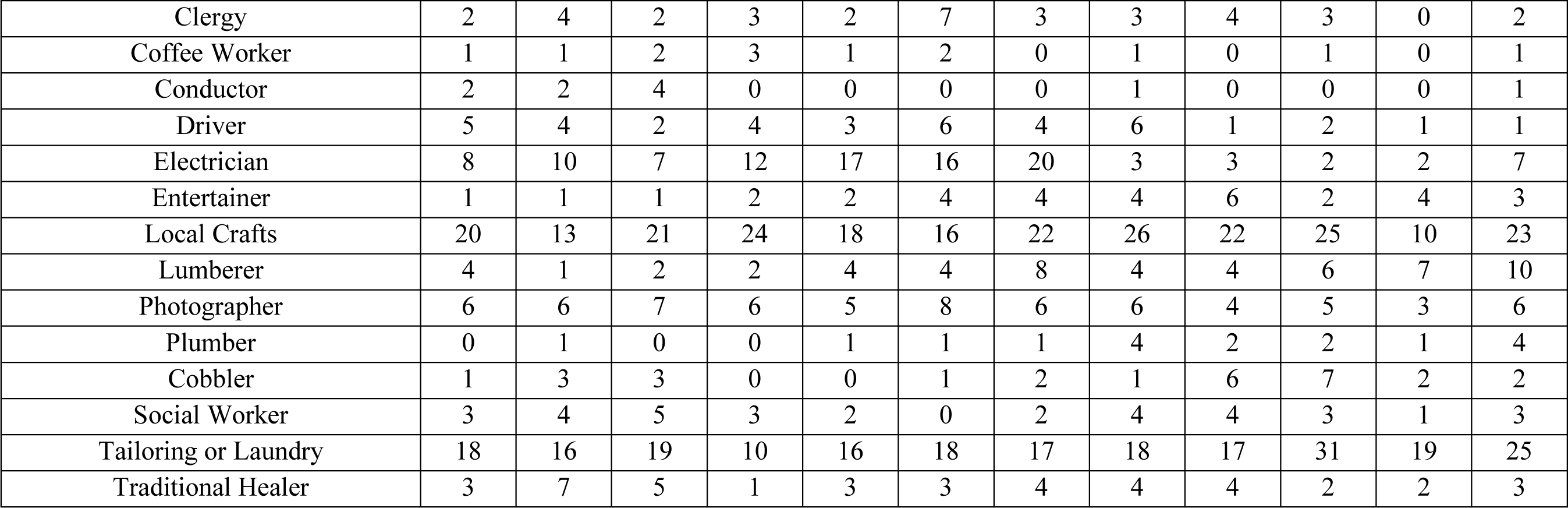
Self-reported primary occupations by male RCCS study participants at each study visit

**Supplemental Table 1b.**
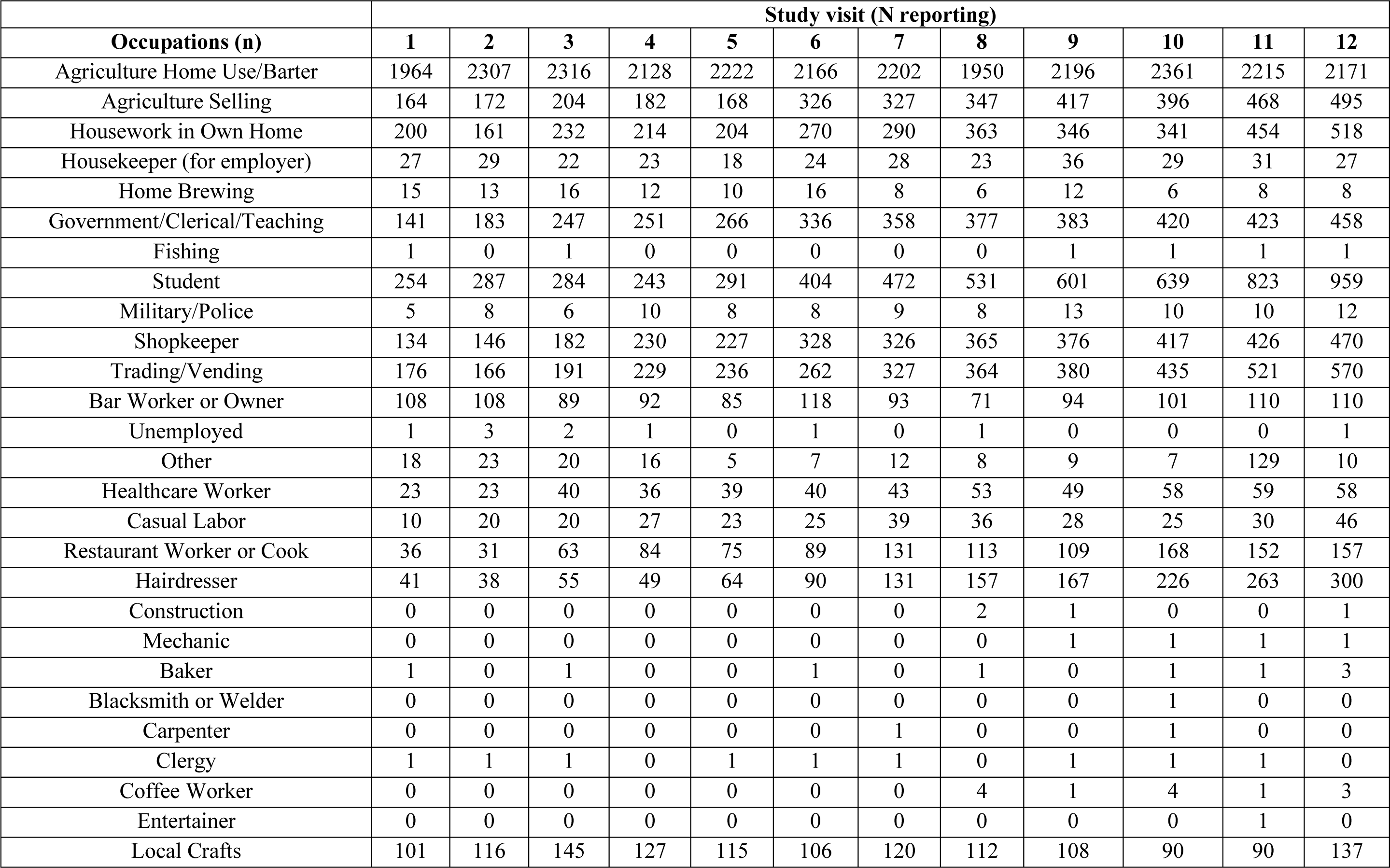
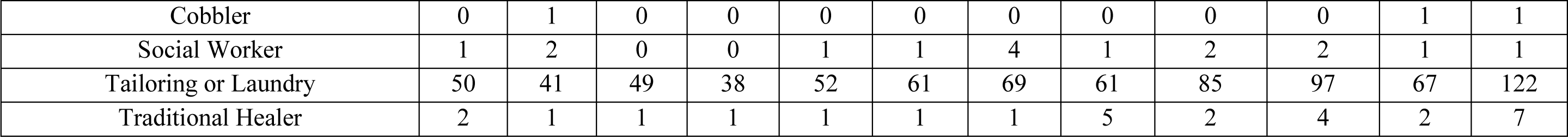
Self-reported primary occupations by female RCCS study participants at each study visit

**Supplemental Table 2.** Recategorization of 41 self-reported primary occupations into occupational sub-groups

| **Final Subgroup** | **Original Category** |
| --- | --- |
| Agriculture | Agriculture home use/barter |
|  | Agriculture selling |
|  | Coffee worker |
| Bar/restaurant work | Bar worker or owner |
|  | Restaurant worker or cook |
|  | Baker |
| Butcher | Butcher |
| Casual labor/unemployed | Home Brewing |
|  | Unemployed |
|  | Casual Labor |
|  | Blacksmith or welder |
|  | Carpenter |
| Construction | Construction |
| Civil service | Government/clerical/teaching |
|  | Military/police |
|  | Healthcare worker |
|  | Social worker |
| Hairdressing | Hairdresser |
| Housekeeping | Housework in own home |
|  | Housekeeper (for employer) |
| Local crafts | Local crafts |
| Mechanic | Mechanic |
| Other | Clergy |
|  | Fishing |
|  | Entertainer |
|  | Cobbler |
|  | Traditional healer |
| Student | Other |
| Tailoring/laundry | Student |
| Trading | Tailoring or laundry |
|  | Shopkeeper |
| Transportation | Trader |
|  | Trucker |
|  | Boda Boda |
|  | Bus conductor |
|  | Driver |

**Supplemental Table 3a.** Number of male observations at each study visit by primary occupational subgroup

| Occupational subgroup | Study Visit<br>N = 44,286<sup>a</sup> |  |  |  |  |  |  |  |  |  |  |  | Median number of observations across all visits |
| --- | --- | --- | --- | --- | --- | --- | --- | --- | --- | --- | --- | --- | --- |
|  | 1 | 2 | 3 | 4 | 5 | 6 | 7 | 8 | 9 | 10 | 11 | 12 |  |
| Agriculture | 975 | 1159 | 1237 | 1021 | 1034 | 1189 | 1197 | 1199 | 1458 | 1438 | 1543 | 1538 | 1198.5 |
| Bar/restaurant worker<sup>b</sup> | 11 | 11 | 10 | 24 | 23 | 21 | 23 | 26 | 21 | 34 | 29 | 32 | 23 |
| Butcher<sup>b</sup> | 8 | 15 | 23 | 26 | 23 | 21 | 36 | 33 | 23 | 32 | 28 | 33 | 24.5 |
| Casual labor | 66 | 84 | 114 | 104 | 96 | 104 | 122 | 79 | 96 | 117 | 110 | 132 | 104 |
| Civil service | 221 | 260 | 321 | 311 | 317 | 320 | 334 | 314 | 349 | 408 | 382 | 429 | 320.5 |
| Construction | 285 | 284 | 337 | 340 | 333 | 333 | 356 | 385 | 367 | 474 | 460 | 500 | 348 |
| Hairdressing<sup>b</sup> | 7 | 12 | 23 | 24 | 29 | 33 | 34 | 41 | 43 | 52 | 57 | 68 | 33.5 |
| Housekeeping<sup>b</sup> | 6 | 7 | 9 | 9 | 13 | 6 | 11 | 12 | 9 | 12 | 21 | 23 | 10 |
| Local crafts<sup>b</sup> | 20 | 13 | 21 | 24 | 18 | 16 | 22 | 26 | 22 | 25 | 10 | 23 | 21.5 |
| Mechanic | 62 | 60 | 85 | 70 | 64 | 97 | 119 | 158 | 191 | 233 | 247 | 302 | 108 |
| Student | 319 | 351 | 388 | 343 | 333 | 432 | 543 | 582 | 726 | 851 | 988 | 1178 | 488 |
| Tailoring/laundry<sup>b</sup> | 18 | 16 | 19 | 10 | 16 | 18 | 17 | 18 | 17 | 31 | 19 | 25 | 18 |
| Trading | 401 | 445 | 418 | 407 | 468 | 494 | 578 | 610 | 625 | 723 | 698 | 756 | 536.5 |
| Transportation | 49 | 59 | 72 | 95 | 92 | 141 | 169 | 198 | 214 | 259 | 244 | 252 | 155 |
| Other<sup>c</sup> | 70 | 75 | 73 | 54 | 64 | 70 | 52 | 43 | 63 | 57 | 186 | 67 | 65.5 |
| Total | 2518 | 2851 | 3150 | 2862 | 2923 | 3295 | 3613 | 3724 | 4224 | 4746 | 5022 | 5358 | 3454 |

<sup>a</sup>14,753 unique individuals
<sup>b</sup>Occupations were dropped from final analysis if median number of observations across all study visits < 50
<sup>c</sup>Category is a composite of multiple small categories that do not fit in any of the main categories and was dropped from main analysis
n – Number of observations by visit in each occupational category
N – Total number of observations during study period

**Supplemental Table 3b.** Number of female observations in each primary occupational subgroup at each study visit

| Occupational subgroup | Study Visit<br>N = 58,473<sup>a</sup> |  |  |  |  |  |  |  |  |  |  |  | Median number of observations across all visits |
| --- | --- | --- | --- | --- | --- | --- | --- | --- | --- | --- | --- | --- | --- |
|  | 1 | 2 | 3 | 4 | 5 | 6 | 7 | 8 | 9 | 10 | 11 | 12 |  |
| Agriculture | 2128 | 2479 | 2520 | 2310 | 2390 | 2492 | 2529 | 2301 | 2614 | 2761 | 2684 | 2669 | 2506 |
| Bar/restaurant worker | 144 | 139 | 152 | 176 | 160 | 207 | 224 | 184 | 203 | 269 | 262 | 267 | 193.5 |
| Butcher<sup>b</sup> | 0 | 0 | 0 | 0 | 0 | 0 | 0 | 0 | 0 | 0 | 0 | 0 | 0 |
| Casual Labor<sup>b</sup> | 11 | 23 | 22 | 28 | 23 | 26 | 39 | 37 | 28 | 25 | 30 | 47 | 27 |
| Civil service | 170 | 216 | 293 | 297 | 314 | 385 | 414 | 439 | 447 | 490 | 493 | 529 | 399.5 |
| Construction<sup>b</sup> | 0 | 0 | 0 | 0 | 0 | 0 | 1 | 2 | 1 | 2 | 0 | 1 | 1 |
| Hairdressing | 41 | 38 | 55 | 49 | 64 | 90 | 131 | 157 | 167 | 226 | 263 | 300 | 110.5 |
| Housekeeping | 227 | 190 | 254 | 237 | 222 | 294 | 318 | 386 | 382 | 370 | 485 | 545 | 306.5 |
| Local Crafts | 101 | 116 | 145 | 127 | 115 | 106 | 120 | 112 | 108 | 90 | 90 | 137 | 113.5 |
| Mechanic<sup>b</sup> | 0 | 0 | 0 | 0 | 0 | 0 | 0 | 0 | 1 | 1 | 1 | 1 | 1 |
| Student | 254 | 287 | 284 | 243 | 291 | 404 | 472 | 531 | 601 | 639 | 823 | 959 | 438.5 |
| Tailoring/Laundry | 50 | 41 | 49 | 38 | 52 | 61 | 69 | 61 | 85 | 97 | 67 | 122 | 61 |
| Trading | 325 | 325 | 389 | 471 | 473 | 606 | 661 | 735 | 768 | 858 | 955 | 1048 | 633.5 |
| Transportation | 0 | 0 | 0 | 0 | 0 | 0 | 0 | 0 | 0 | 0 | 0 | 0 | 0 |
| Other<sup>b</sup> | 23 | 26 | 24 | 17 | 7 | 10 | 14 | 14 | 13 | 14 | 136 | 22 | 15.5 |
| **Total** | **3474** | **3880** | **4187** | **3993** | **4111** | **4681** | **4992** | **4959** | **5418** | **5842** | **6289** | **6647** | **4820** |

<sup>a</sup>19,113 unique individuals
<sup>b</sup>Occupations were dropped from final analysis if median number of observations across all study visits < 50
n – Number of observations by visit in each occupational category
N – Total number of observations during study period

## Notes

### Funding Statement

This study was funded by NIAID, NIMH, NICHD, and the US CDC.

### Author Declarations

The Research and Ethics Committee of the Uganda Virus Research Institute, the Ugandan Council of Science and Technology, and the Johns Hopkins School of Medicine Institutional Review Board gave ethical approval for this work.

